# Device measured sedentary behaviour, sleep, light and moderate-vigorous physical activity and cardio-metabolic health: A compositional individual participant data analysis in the ProPASS consortium

**DOI:** 10.1101/2023.08.01.23293499

**Authors:** JM Blodgett, MN Ahmadi, AJ Atkin, S Chastin, HW Chan, K Suorsa, EA Bakker, P Hettiarcachchi, PJ Johansson, LB Sherar, V Rangul, RM Pulsford, G Mishra, TMH Eijsvogel, S Stenholm, AD Hughes, AM Teixeira-Pinto, U Eklund, IM Lee, ProPASS collaboration, A Holtermann, A Koster, E Stamatakis, M Hamer

## Abstract

**Background/Aims:** Physical inactivity, sedentary behaviour (SB) and inadequate sleep are key behavioural risk factors of cardiometabolic diseases; each is mainly considered in isolation. The study aim was to investigate associations of five movement behaviour compositions with adiposity and cardiometabolic biomarkers.

**Methods:** Cross-sectional data from 15,246 participants from the Prospective Physical Activity, Sitting and Sleep consortium (ProPASS) were analysed. Time spent in sleep, SB, standing, light-intensity physical activity (LIPA) and moderate-vigorous physical activity (MVPA) made up the composition. Outcomes included BMI, waist circumference, HDL cholesterol, total:HDL cholesterol ratio, triglycerides and HbA1c. Compositional linear regression examined associations between compositions and each outcome, including modelling reallocation of time between behaviours.

**Results:** The average daily composition of the sample(age:53.7±9.7years; 54.7%female) was 7.7hrs sleeping,10.4hrs sedentary,3.1hrs standing,1.5hrs LIPA and 1.3hrs MVPA. A greater proportion of MVPA time and smaller proportion of SB time was associated with better outcomes. Reallocating time from SB,standing,LIPA or sleep into MVPA had the largest theoretical improvement across all outcomes. For example, replacing 30min of SB, sleep, standing or LIPA with MVPA was associated with -0.63 (95%CI -0.48,-0.79), -0.43 (-0.25,-0.59), -0.40 (-0.25,-0.56) and -0.15 (0.05,-0.34)kg/m^2^ lower BMI, respectively. A larger proportion of standing time was beneficial for outcomes; sleep had a detrimental association when replacing LIPA or MVPA and positive association when replacing SB. The minimal displacement into MVPA for improved cardiometabolic health ranged from 3.8 (HbA1c) to 12.7 (triglycerides) min/day.

**Conclusions:** Compositional data analyses revealed a distinct hierarchy of behaviours. MVPA demonstrated the strongest, most time-efficient protective associations with cardiometabolic outcomes. Theoretical benefits from reallocating SB into sleep, standing or LIPA required substantial changes in daily activity.

## INTRODUCTION

Cardiometabolic diseases - including cardiovascular disease (CVD), obesity and diabetes mellitus - are the leading cause of mortality worldwide^1^. The global burden of these diseases has risen over the past three decades, with annual CVD-related deaths increasing from 12.1 to 18.6 million, while diabetes-related deaths have doubled to 1.25 million^2, 3^. Concerningly, these trends are forecasted to continue^4, 5^. Positive engagement in health behaviours, such as physical activity, reducing sedentary behaviour, and ensuring sufficient quality and quantity of sleep, can help prevent cardiometabolic disease^1, 6^, yet are largely underutilised.

Regular moderate-vigorous physical activity (MVPA) has established cardiometabolic benefits via direct inflammatory, metabolic or cardiovascular mechanisms^7, 8^. However, the effects of light intensity physical activity (LIPA) are less clear^9^. This may be due to poor ascertainment of LIPA using self-reported questionnaires^10^ or threshold-based approaches of hip or wrist-based accelerometery which fail to distinguish between standing and subtle ambulatory activities^11^. There is a strong argument against classifying passive standing as LIPA, given the very low energy expenditure involved^12^. Finally, there is consistent evidence of associations between sedentary behaviour (SB) and increased cardiometabolic disease risk^13^, while the role of sleep is less clear^14^.

Time spent in these daily movement behaviours (sleep, SB, standing, LIPA, MVPA) form a 24-hour composition, with any change in one behaviour resulting in a corresponding increase or decrease in another. However, controlled exercise trials and observational studies have mainly examined each behaviour in isolation^9, 14, 15^. Assumptions that these behaviours are independent and that the 24-hr day is infinite can lead to potentially imprecise estimates that cannot be translated to real-world interventions or guidelines. Treating these data as a complete 24-hour day using compositional data analysis can overcome this limitation^16^. Previous evidence of movement compositions have suggested that more time in MVPA and less time in SB are associated with favourable health outcomes^17–19^. However, these studies have largely relied upon small sample sizes, considered compositions with awake time only or incorporated self-reported sleep measures, and were unable to differentiate between sedentary and standing activity (i.e. due to wrist or hip worn accelerometers).

The majority of current public health guidelines (i.e. WHO, USA, UK) focus solely on physical activity and sedentary behaviour^20, 21^. There is a clear need for better empirical evidence to support “24-hour” guidelines^22^ and encompass recommendations on daily sleep, SB and activity intensity volume. The Prospective Physical Activity, Sitting, and Sleep consortium (ProPASS) resource^23^ overcomes major limitations of previous literature^17–19^ by using harmonized individual-level data from six studies with thigh-worn accelerometery and a unified approach to derive 24-hour movement behaviours. Our aim was to examine the associations between compositions of movement behaviours (defined as time spent in sleep, SB, standing, LIPA, MVPA) and six cardiometabolic outcomes. Using the mean sample behavioural profile, we also estimated the impact of reallocating time from one behaviour to another.

## METHODS

### Sample

ProPASS is an international research collaboration platform consisting of observational cohort studies with thigh-worn accelerometry^23^. We used cross-sectional data from six participating studies: The Maastricht Study (TMS; Netherlands, n=7515)^24^, the 1970 British Birth Cohort Study (BCS70; United Kingdom, n=5263)^25^, the Australian Longitudinal Study on Women’s Health (ALSWH; Australia, n=950)^26^, the Danish PHysical ACTivity cohort with Objective measurements cohort (DPhacto; Denmark, n=780)^27^, the Nijmegen Exercise Study (NES; Netherlands, n=537)^28^and the Finnish Retirement and Aging Study (FIREA; Finland, n=253)^29^. Ethical approval and informed consent were provided for each cohort; study details are available elsewhere^23–29^. Device-measured activity data were available for 15,271 participants.

### Movement behaviours

All cohorts collected movement behaviour data using a 7-day, 24-hour/day thigh-worn accelerometer protocol; four studies used ActivPAL3/4 devices (BCS70, TMS, ALSWH, NES;), one used Axivity devices (FIREA) and one used ActiGraph devices (DPhacto). Raw accelerometer data was centrally processed using previously validated software, ActiPASS v 1.32, which implements algorithms for non-wear, sleep detection and validated activity across different thigh-worn accelerometer brands^11, 30–33^. ActiPASS identifies behaviours in 2-second windows with a 50% overlap, resulting in a resolution of 1-second epochs. We classified five movement behaviours: sleep, SB (sitting or lying episodes outside of sleep intervals), standing, LIPA (ambulatory movement without purposeful walking, walking with cadence <100steps/minutes) and MVPA (running, cycling, inclined stepping, walking with cadence ≥100steps/minute)^11, 30–34^. Participants with at least one valid wear day (≥20 hours of wear/day)^25^, ≥1 period of walking detection and >0 minutes of sleep were included in analyses. Time spent in each behaviour was calculated as average minutes/day.

### Cardiometabolic outcomes

Two markers of adiposity were assessed by trained nurses or researchers during home or clinic-based visits: *body mass index* (BMI, kg/m^2^; calculated from height and weight) and *waist circumference* (cm). Cardiometabolic blood biomarkers were measured in five studies (not available in DPhacto) and included: *high-density lipoprotein cholesterol* (HDL; mmol/L), *total:HDL cholesterol ratio*, *triglycerides* (mmol/L) and *HbA1c* (glycated hemoglobin, mmol/mol; measured in ALSWH, BCS70 and TMS only). Full details of ascertainment of outcomes by study, including assay details, are provided in Supplementary Tables 1 and 2.

### Covariates

Covariates were selected a priori based on data availability and known associations with movement behaviours and cardiovascular outcomes^17–19^. The following covariates were collected in all cohorts: *age* (years), *sex* (male, female), *smoking status* (non-smoker, current smoker), *alcohol consumption* (tertiles based on self-reported weekly consumption), *self-rated health* (five-point Likert scale), *lipid-modifying, hypertensive or glucose-lowering medications* (yes, no) and *history of cardiovascular disease* (CVD; yes, no). Additionally, a subset of cohorts collected data on *mobility limitations* (n=4 cohorts; continuous score from 0 to 100 of the SF-36 10-item physical function subscale, where 0 indicates poor mobility and 100 indicates no mobility problems), o*ccupational class* (n=5 cohorts; not working, low, intermediate, high occupational class) and *education* (n=4 cohorts; none or lower than high school, high school qualifications/typically attained at age 16y, further education qualifications/typically attained at age 16-18y, university degree and higher/typically 18+y). Full details of ascertainment and subsequent harmonisation of covariates in each cohort are provided in Supplementary Table 1.

### Statistical analyses

We define a composition as the average daily time spent in each of SB, sleep, standing, LIPA and MVPA behaviours. First, average daily times are normalised such that the sum of all behaviours is equivalent to 1440 minutes (24 hours) to account for any non-wear time. The 24-hour time composition is then expressed as a set of four isometric log-ratio (*<u>ilr</u>*) coordinates ^16, 35^, capturing information and variability of the relative time spent in each of the five behaviours. Therefore, we used the following set of *<u>ilr</u>* coordinates to capture time spent in all five behaviours: 1) SB compared to sleep, standing, LIPA and MVPA; 2) sleep compared to standing, LIPA and MVPA; 3) standing compared to LIPA and MVPA; 4) LIPA compared to MVPA. Inclusion of all four coordinates in a single regression model allows the relation between all behaviours to be captured. We pivoted the data to create five sets of coordinates; each set permits investigation of the first coordinate (i.e., the behaviour of interest relative to time spent in the other four behaviours)^35^.

Using these coordinates, we conducted a one-stage individual participant meta-analysis using linear regression for each outcome. Coefficients indicate the change in outcome (e.g., kg/m^2^ or mmol/L) for each 1-unit *ilr* increase. We tested for sex-interactions before building models in two stages: 1) adjusted for sex, age and cohort; 2) adjusted for sex, age, cohort, smoking, alcohol, self-reported health, medications, and CVD history. Due to cohort-specific missing data, sex-age-cohort adjusted models were examined in both the maximal available sample and those with complete covariate data and differences in movement behaviours and outcomes were compared between subsamples. Sensitivity analyses further considered models adjusted for education, mobility limitations and occupational class in the three cohorts with data on all three additional covariates (ALSWH, BCS70, TMS). Differences in movement behaviours and outcomes were examined across all three sample sizes. To provide results ready for translation to behavioural interventions, we conducted isotemporal substitution to model how reallocation of time from one behaviour to another – based on the mean 24-hour behavioural profile –impacted each outcome^36, 37^ in sex-age-cohort adjusted models. To enable comparisons with existing literature^17^, we repeated regression and reallocation plots using a four-part composition of SB, sleep, MVPA and LIPA, where standing was included as LIPA. All analyses were performed in RStudio using the *cCompositions*, *robCompositions* and *zCompositions* packages.

## RESULTS

### Sample description

Of 15,271 participants with valid accelerometer data on all five behaviours, 15,253 (99.9%) had data on at least one outcome. Table 1 provides descriptive characteristics of the sample for all movement behaviours, outcomes and covariates. Briefly, 54.7% (n=8,341) of the sample were female, with a mean age of 53.7yrs ±9.7 (range: 18-87). The majority of the sample were non-smokers (85.4%), self-rated their health as good or better (87.8%), were not taking take lipid-modifying, hypertensive or glucose-lowering medications (70.1%) and had no history of CVD (90.2%). Average daily wear time across the wear period was 22.8hrs

**Table 1.** Descriptive characteristics in maximal available sample (n=15,253)

| **OUTCOMES mean±SD** |  | **Full sample**<br>**(n=15,243)** | **Females**<br>**(n=8,341; 54.7%)** | **Males**<br>**(n=6,912; 45.3%)** |
| --- | --- | --- | --- | --- |
| BMI (kg/m<sup>2</sup>) |  | 27.0 ±4.9 | 26.7 ±5.4 | 27.4 ±4.3 |
| Waist circumference (cm) |  | 94.1 ±13.9 | 89.2 ±13.2 | 100.2 ±12.1 |
| HDL cholesterol (mmol/L) |  | 1.57 ±0.46 | 1.7 ±0.5 | 1.4 ±0.4 |
| HDL: total cholesterol ratio |  | 3.64 ±1.23 | 3.3 ±1.0 | 4.0 ±1.3 |
| HbA1c (mmol/mol) |  | 38.0 ±8.7 | 36.7 ±7.3 | 39.4 ±9.8 |
| Triglycerides (mmol/L) |  | 1.48 ±1.04 | 1.3 ±0.8 | 1.7 ±1.2 |
| **MOVEMENT BEHAVIOR COMPOSITION \* (hrs/day; % of day)** |  |  |  |  |
| Sleep |  | 7.7 31.9% | 7.9 32.8% | 7.4 30.9% |
| Sedentary behavior |  | 10.4 43.2% | 9.9 42.3% | 10.9 45.4% |
| Standing |  | 3.1 13.0% | 3.3 13.9% | 2.9 11.9% |
| LIPA |  | 1.5 6.4% | 1.5 6.3% | 1.6 6.5% |
| MVPA |  | 1.3 5.5% | 1.4 5.7% | 1.3 5.3% |
| **MAIN ANALYSES COVARIATES mean±SD or n(%)** |  |  |  |  |
| **Age (years)** |  | 53.7 ±9.7 | 52.7 ±9.1 | 55.1 ±10.2 |
| **Cohort** | TMS | 7,515 (49.3) | 3,790 (45.4) | 3,725 (53.9) |
|  | BCS70 | 5,236 (34.3) | 2,797 (33.5) | 2,439 (35.3) |
|  | ALSWH | 941 (6.1) | 941 (11.3) | 0 (0) |
|  | DPhacto | 777 (5.1) | 359 (4.3) | 412 (6.0) |
|  | NES | 537 (3.5) | 244 (2.9) | 293 (4.2) |
|  | FIREA | 253 (1.7) | 210 (2.5) | 43 (0.6) |
| **Smoking status** | Non-smoker | 12,953 (85.4) | 7,205 (86.8) | 5,748 (83.7) |
|  | Current smoker | 2,211 (14.6) | 1,093 (13.2) | 1,118 (16.3) |
| **Alcohol consumption** | Tertile 1 (low) | 4,463 (33.8) | 3,058 (42.6) | 1,405 (23.3) |
|  | Tertile 2 | 4,514 (34.2) | 2,529 (35.2) | 1,985 (32.9) |
|  | Tertile 3 (high) | 4,231 (32.0) | 1,591 (22.2) | 2,640 (43.8) |
| **Self-reported health†** | Excellent | 1,849 (12.3) | 1,102 (13.4) | 747 (11.0) |
|  | Very good | 4,905 (32.7) | 2,701 (32.9) | 2,204 (32.4) |
|  | Good | 6,329 (42.2) | 3,337 (40.6) | 2,992 (44.1) |
|  | Fair | 1,634 (10.9) | 908 (11.1) | 726 (10.7) |
|  | Poor | 287 (1.9) | 164 (2.0) | 123 (1.8) |
| **Medication** (lipid-modifying, hypertensive or glucose-lowering) |  | 4,333 (29.9) | 1,859 23.3) | 2,474 (38.1) |
| **History of CVD** |  | 1,486 (9.8) | 615 7.4) | 871 (12.7) |
| **SUPPLEMENTARY ANALYSES COVARIATES ‡ mean(SD) or n(%)** |  |  |  |  |
| **Physical function (SF-36)** |  | 87.2 ±18.8 | 86.5 ±18.9) | 88.0 ±18.6 |
| **Occupational class** | Not working | 3,850 (29.3) | 2,014 (28.1) | 1,836 (30.6) |
|  | Low | 2,152 (16.4) | 1,023 (14.3) | 1,129 (18.8) |
|  | Intermediate | 3,645 (27.7) | 1,927 (26.9) | 1,718 (28.7) |
|  | High | 3,502 (26.6) | 2,192 (30.6) | 1,310 (21.9) |
| **Education** | None or less than high school | 1,666 (11.9) | 764 (9.9) | 902 (14.2) |
|  | High school (~16y) | 3,908 (27.8) | 2,209 (28.7) | 1,699 (26.7) |
|  | Further education (~16-18y) | 5,399 (38.4) | 2,955 (38.4) | 2,444 (38.4) |
|  | University degree or higher | 3,080 (21.9) | 1,760 (22.9) | 1,320 (20.7) |

\* Re-scaled to a 24-hour day to create the composition
† Note: response terminology differs slightly by cohort, given translation of original question (see Supplementary Table 1)
‡ Covariates available in restricted cohorts only (see Supplementary Table 1)
**ALSWH** Australian Longitudinal Study of Women's Health; **BMI** Body Mass Index; **BCS70** 1970 British Cohort Study; **CVD** cardiovascular disease; **DPhacto** Danish PHysical ACTivity cohort with Objective measurements; **FIREA** Finnish Retirement and Aging Study; **HDL** High Density Lipoprotein; **LIPA** Light-intensity Physical Activity; **MVPA** Moderate-Vigorous intensity Physical Activity; **NES** Nijmegen Exercise Study; **SD** standard deviation; **SF-36** Short-Form 36; **TMS** The Maastricht Study

±1.8. The mean composition of the full sample, defined as the average time spent in each behaviour normalised to a 24-hour day, was 7.7 hrs sleeping, 10.4 hrs sedentary, 3.1 hrs standing, 1.5 hrs in LIPA and 1.3 hrs in MVPA. Supplementary Figure 1a demonstrates absolute differences in time spent in each movement behaviour by cohort, while Supplementary Figure 1b provides percent differences compared to the overall mean sample composition. Inter-cohort differences were largest for standing, LIPA and MVPA, with comparable time spent sleeping and in SB. The maximal available sample in sex-age-cohort adjusted models ranged from 11,270 (triglycerides; n=9,450 complete cases) to 15,204 (BMI; n=12,166 complete cases).

### Association between movement behaviours and adiposity

A greater proportion of time spent sedentary was associated with higher BMI (Supplemental Table 3); conversely – and in order of size of association – more time engaging in MVPA, LIPA, standing or sleep was associated with lower BMI. Associations were robust to adjustment for all covariates (Models 2-3, Supplemental Tables 3-4). Reallocation of time from any behaviour into MVPA, while holding the others constant, had the largest theoretical reduction in BMI (Figure 1). For example, reallocating 30 minutes of SB, sleep, standing or LIPA into MVPA was associated with -0.63 (95% CI: -0.48, -0.79), - 0.43 (-0.25, -0.59), -0.40 (-0.25, -0.56), or -0.15 (0.05, -0.34) kg/m^2^ lower BMI, respectively. Conversely, reallocating time from LIPA or MVPA into sleep, standing or SB was associated with higher BMI (Figure 1a-b). The minimal daily behavioural change required to observe significant theoretical reductions in BMI was displacement of 7.2 minutes of SB into MVPA.

**Figure 1.**
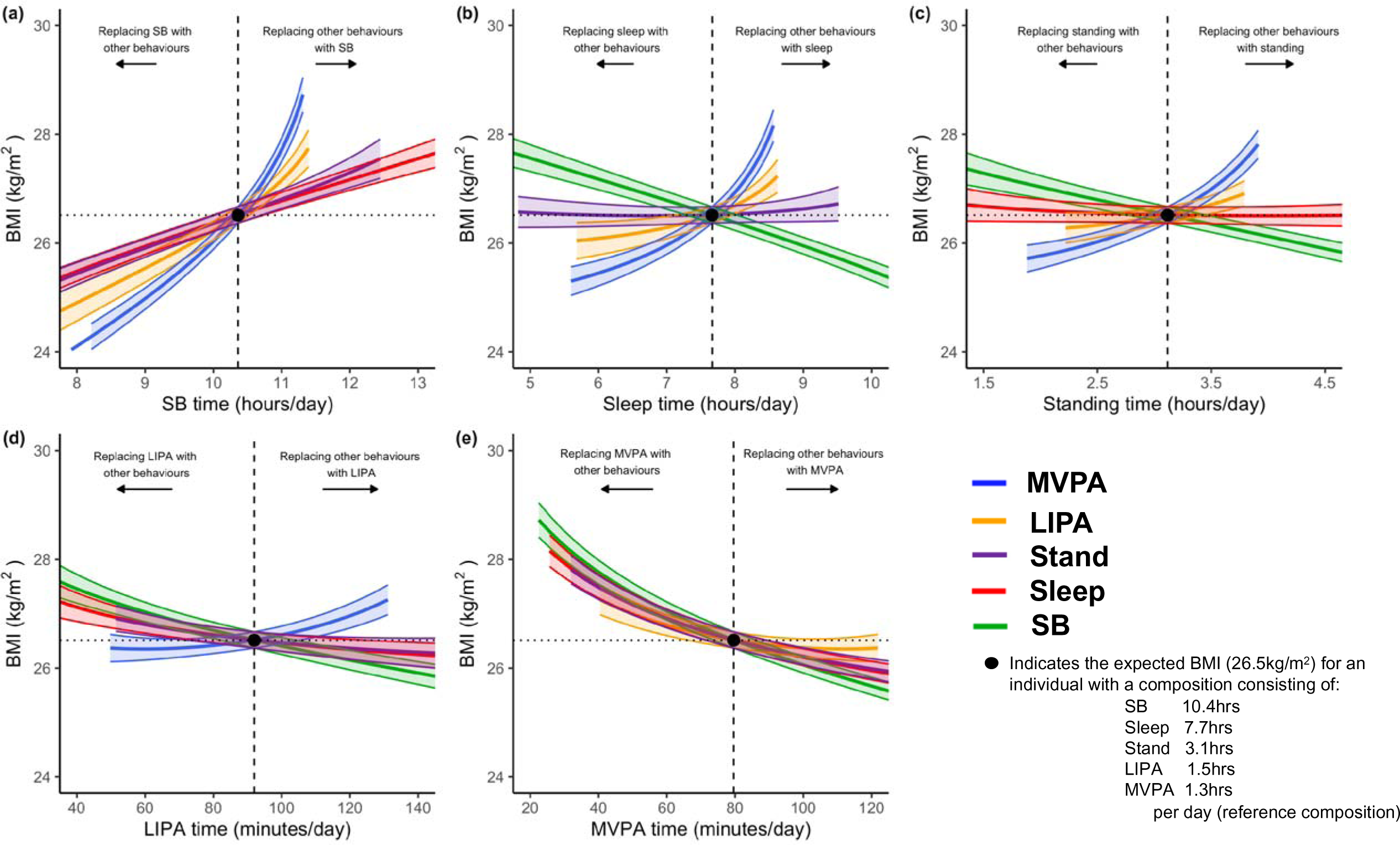
Substitution models (n=15,204) for ***<u>BMI</u>*** for a) sedentary behavior; b) sleep; c) Standing; d) Light Intensity Physical Activity (LIPA); e) Moderate-to-Vigorous Intensity Physical Activity (MVPA). Data to the left of the reference line indicates the predicted change in BMI if a given behavior (e.g. SB in panel a) is replaced by each of the other four behaviors. Data to the right of the reference line indicates the predicted change in BMI if a given behavior (e.g. SB in panel a) replaces each of the other four behaviors. Model adjusted for sex (ref: female), age (ref: 53.7 years; mean-centred) and cohort (ref: Maastricht Study).

Associations were similar for waist circumference across MVPA, standing, sleep and SB (Figure 2). Reallocating 30 minutes of SB, sleep or standing into MVPA was associated with lower waist circumferences of -2.44 (-1.97, -2.78), 1.75 (-1.38, -2.22), and -1.34 (-0.98, -1.78) cm, respectively. Although displacement of LIPA into MVPA remained favourable for waist circumference (30 min: -2.49 (-1.95, -2.94) cm), there was a negative association with waist circumference if time spent in LIPA replaced time spent sleeping or standing (Figure 2d). However, associations were attenuated after adjustment for covariates (Models 2-3, Supplemental Tables 3-4). The minimal behavioural change required to observe statistically significant theoretical reductions in waist circumference was displacement of 5.0 min/day of LIPA into MVPA.

**Figure 2.**
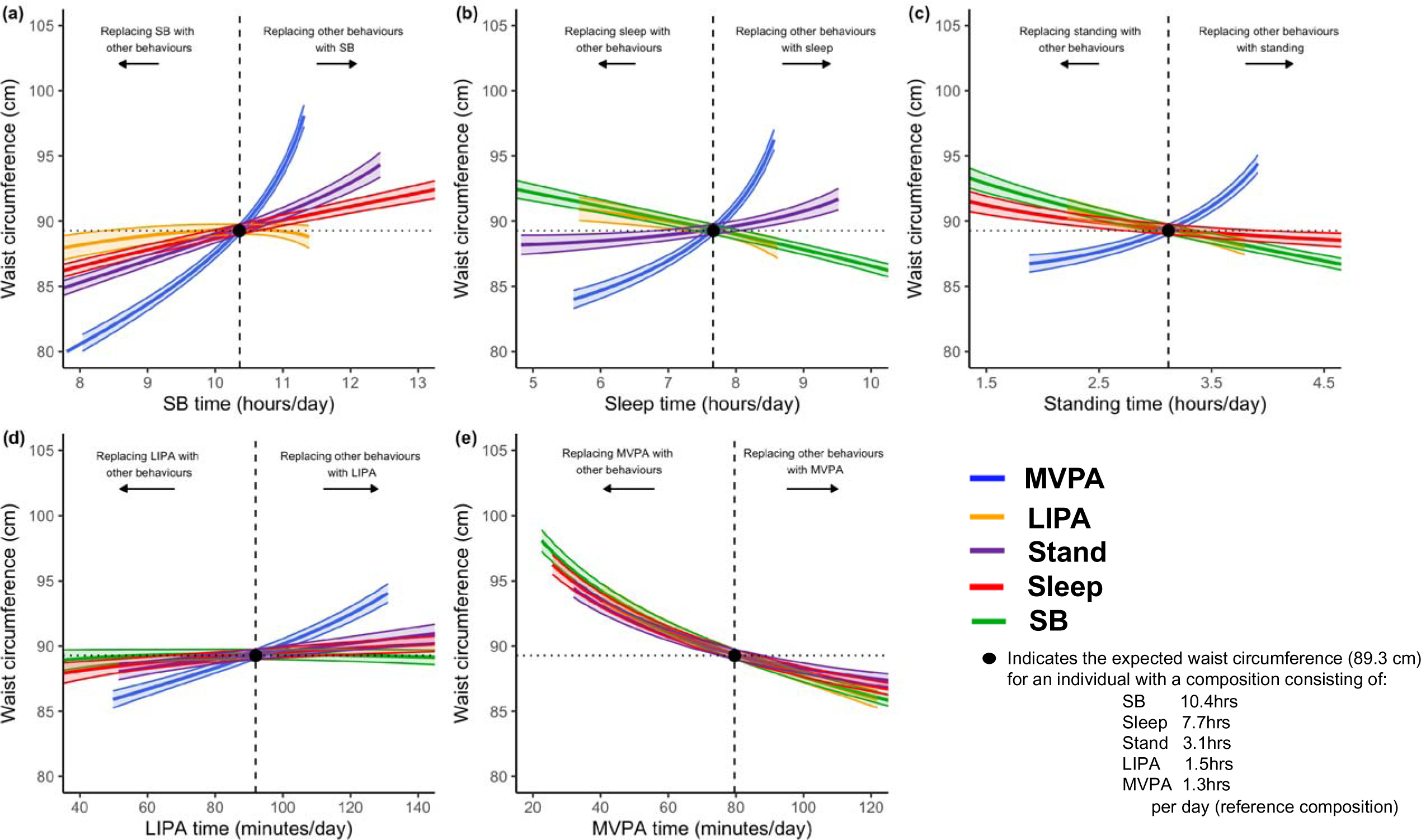
Substitution models (n=14,541) for ***<u>waist circumference</u>*** outcome for a) sedentary behavior; b) sleep; c) Standing; d) Light Intensity Physical Activity (LIPA); e) Moderate to Vigorous Intensity Physical Activity (MVPA). Model adjusted for sex (ref: female), age (ref: 53.7 years; mean-centred) and cohort (ref: Maastricht Study)

### Association between movement behaviours and lipids

A smaller proportion of time in SB and a greater proportion in MVPA was associated with higher HDL cholesterol, lower total:HDL cholesterol ratio and lower triglyceride levels (Supplementary Table 3; Figure 3-5a,e). For example, reallocation models suggested that improvements were observed after as few as 6.0, 8.9 and 12.7 minutes of SB were replaced by MVPA (Figure 3-5e), respectively. Associations remained after adjustment for covariates (Models 2-3, Supplemental Tables 3-4).

**Figure 3.**
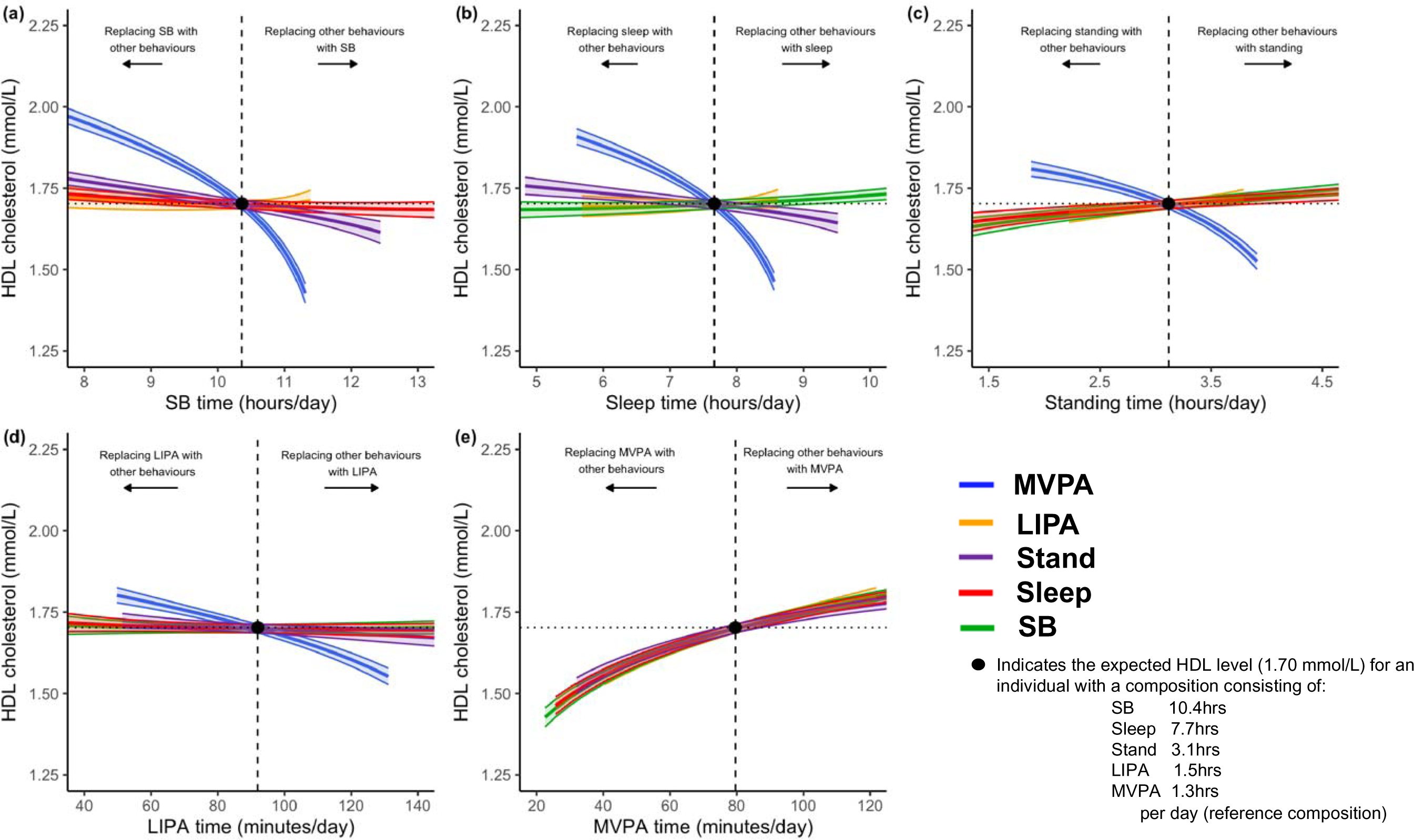
Substitution models (n=13,060) for ***<u>HDL cholesterol</u>*** outcome for a) sedentary behavior; b) sleep; c) Standing; d) Light Intensity Physical Activity (LIPA); e) Moderate to Vigorous Intensity Physical Activity (MVPA). Model adjusted for sex (ref: female), age (ref: 53.7 years; mean-centred) and cohort (ref: Maastricht Study).

**Figure 4.**
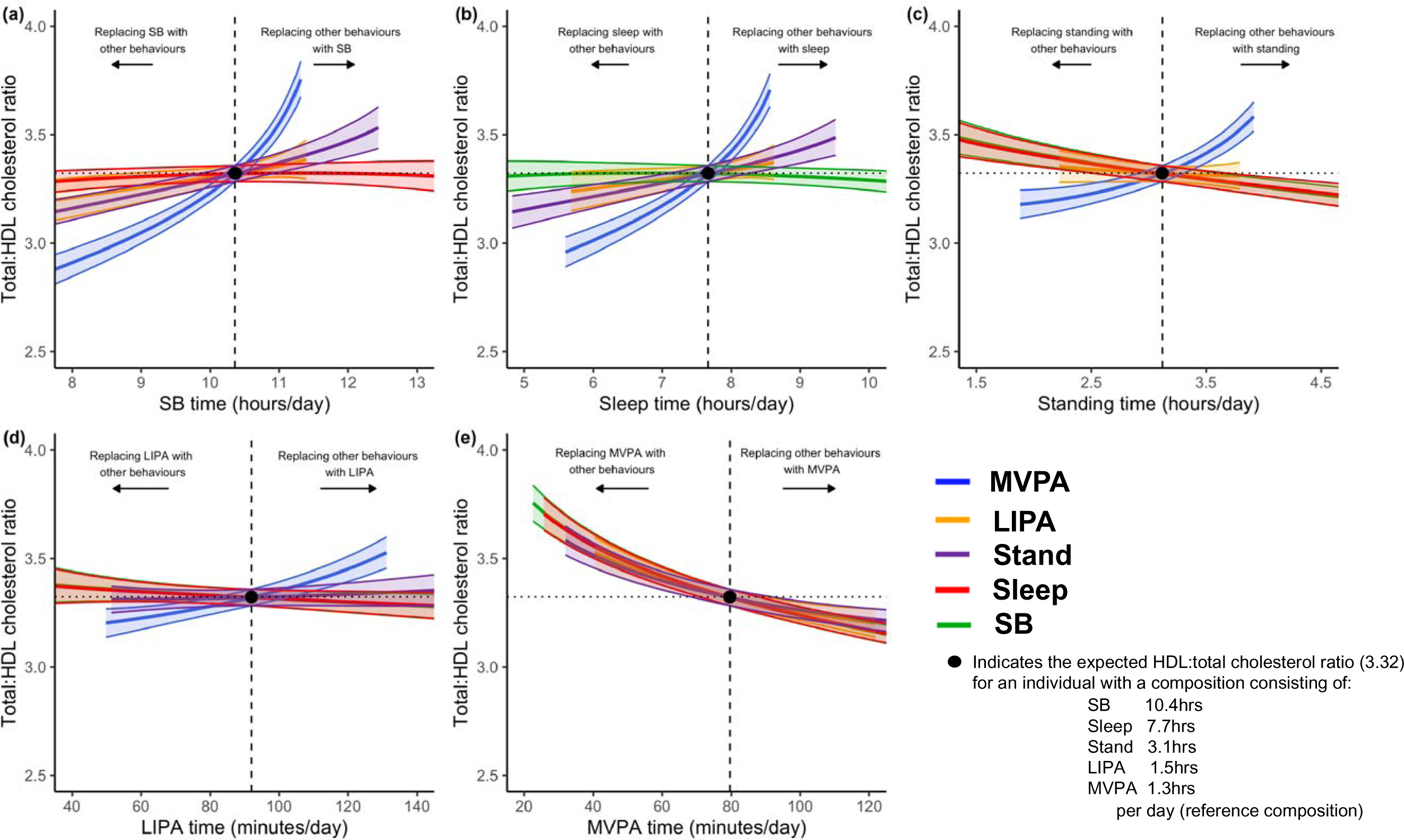
Substitution models (n=13,059) for ***<u>total:HDL cholesterol ratio</u>*** outcome for a) sedentary behavior; b) sleep; c) Standing; d) Light Intensity Physical Activity (LIPA); e) Moderate to Vigorous Intensity Physical Activity (MVPA). Model adjusted for sex (ref: female), age (ref: 53.7 years; mean-centred) and cohort (ref: Maastricht Study).

**Figure 5.**
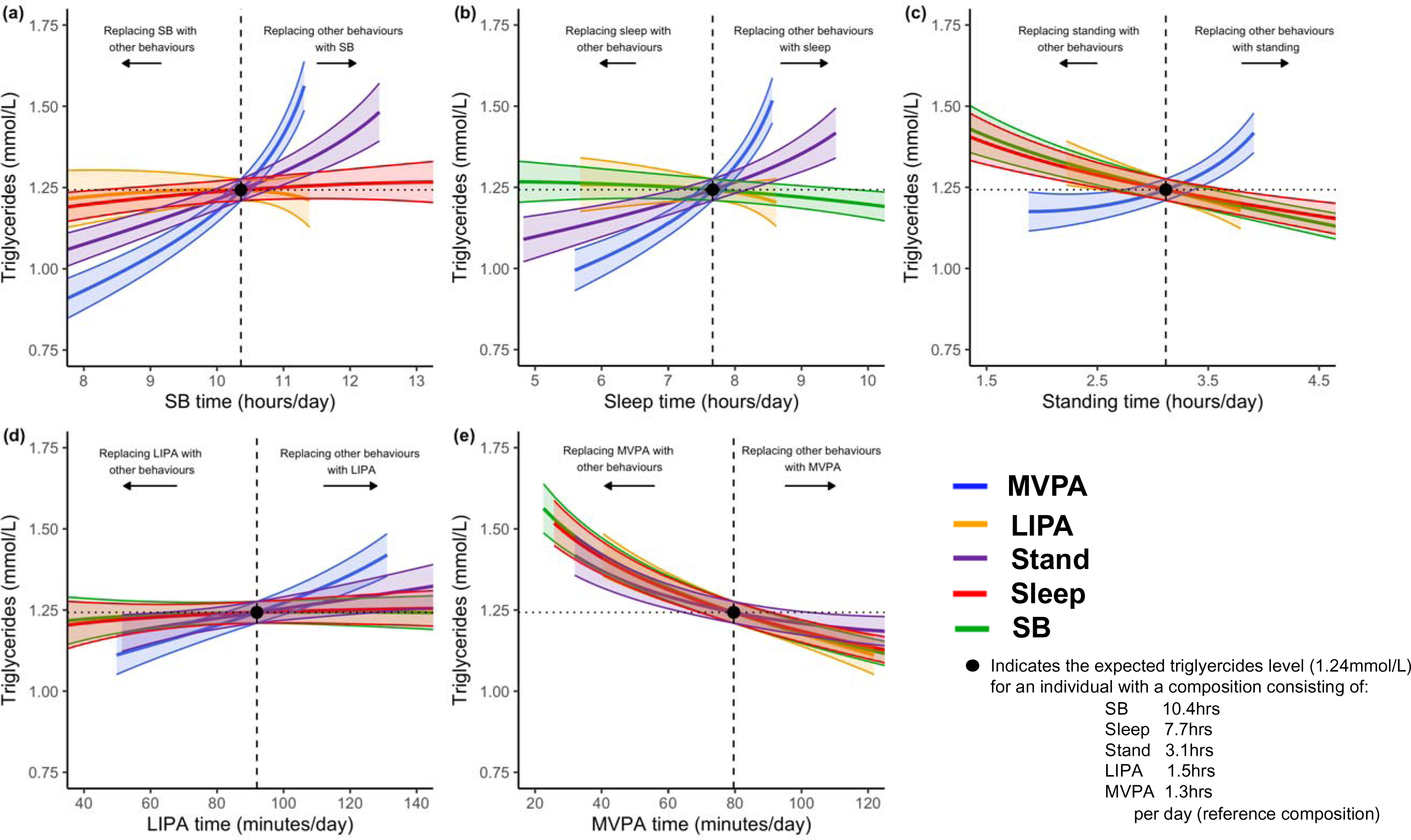
Substitution models (n=12,240) for ***<u>triglycerides</u>*** outcome for a) sedentary behavior; b) sleep; c) Standing; d) Light Intensity Physical Activity (LIPA); e) Moderate to Vigorous Intensity Physical Activity (MVPA). Model adjusted for sex (ref: female), age (ref: 53.7 years; mean-centred) and cohort (ref: Maastricht Study).

Beyond the beneficial impact of reallocating time from LIPA to MVPA, there was little evidence that LIPA displacement was associated with HDL or total:HDL cholesterol ratio (Figures 3-5d, Supplementary Tables 3-4). Conversely, positive associations between a greater proportion of time spent standing and favourable lipid outcomes remained across all outcomes and models. Standing was detrimental when displacing MVPA time, but advantageous when replacing 1+hr sleep or 1.75+hr of SB (Figures 3-5c). Reallocating time between LIPA and standing – in either direction – was negligible for HDL and total:HDL cholesterol ratio, while theoretical reductions in triglycerides level were observed after 39 minutes of LIPA was displaced into standing.

Finally, the role of sleep differed by displaced behaviour and outcome (Figure 3-5b). When sleep displaced MVPA or standing time (Figures 4-6b), there were deleterious associations with all outcomes. For example, replacing 30 minutes of MVPA with sleep was associated with a -0.10 mmol/L (-0.08, -0.12), +0.17 (0.12,0.21) and +0.13 mmol/L (0.08, 0.17) difference in HDL, total:HDL cholesterol ratio and triglycerides. Reallocation between sleep, SB and LIPA was negligible, with a meaningful change in HDL only emerging after ∼1.5 hours of displacement from SB to sleep (Figure 3b).

**Figure 6.**
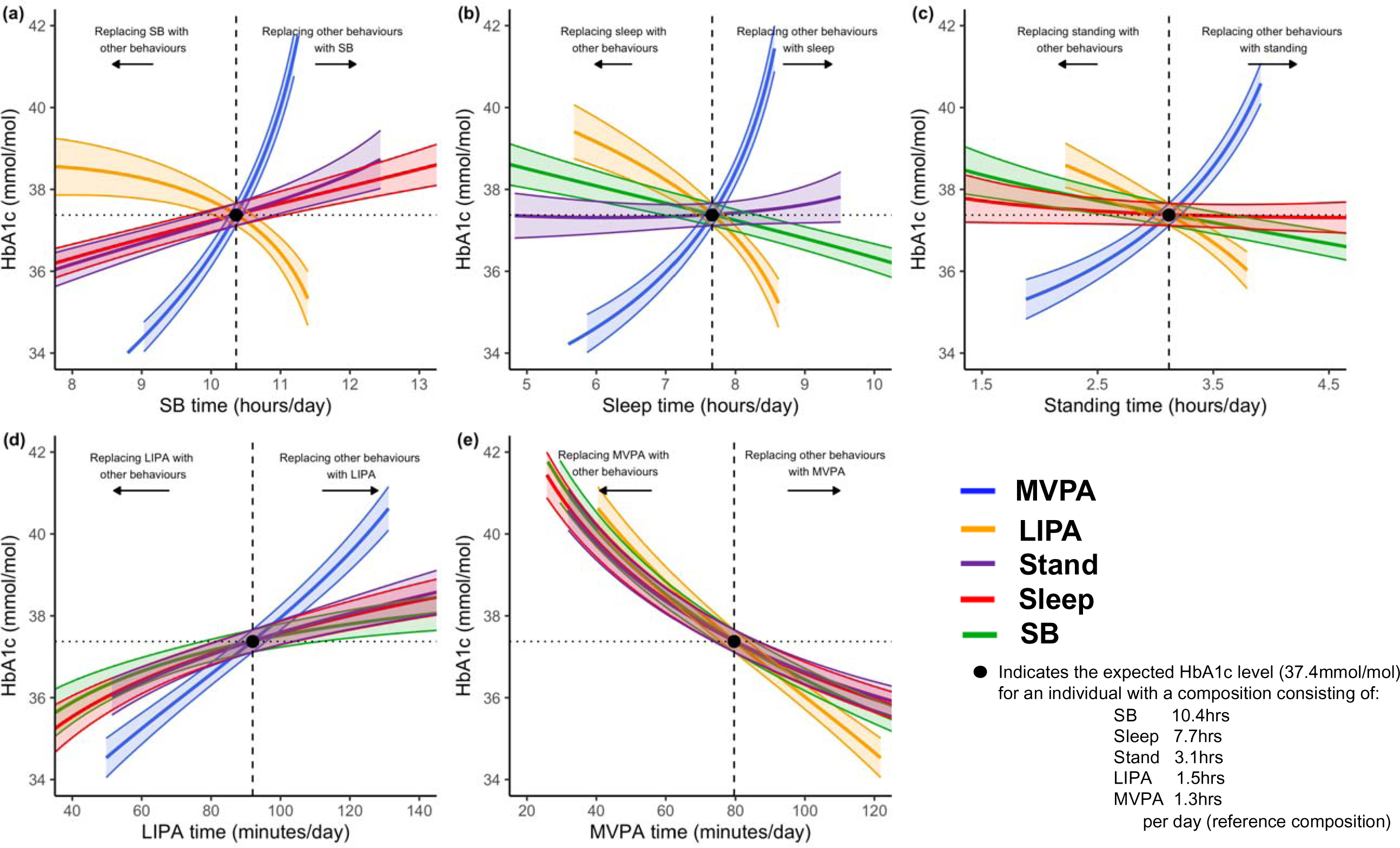
Substitution models (n=11,270) for ***<u>HbA1c</u>*** outcome for a) sedentary behavior; b) sleep; c) Light Intensity Physical Activity (LIPA); d) Moderate to Vigorous Intensity Physical Activity (MVPA). Model adjusted for sex (ref: female), age (ref: 53.7 years; mean-centred) and cohort (ref: Maastricht Study)

### Association between movement behaviours and HbA1c

A greater proportion of time spent in MVPA, standing or sleeping and a smaller proportion of time spent in SB was associated with lower HbA1c. Associations remained after adjustment for covariates (Supplementary Tables 3-4). Relative to other time reallocations, displacement of any other behaviour into MVPA was associated with the most favourable estimates for HbA1c levels (Figure 6). When MVPA replaced 30 minutes spent in SB, sleep, standing or LIPA, we observed lower HbA1c of 1.33(1.06, 1.61), 1.12(0.80, 1.40), 1.04 (0.72, 1.36) and 2.00 (1.63, 2.37) mmol/mol, respectively (Figure 6e).

LIPA was the most deleterious behaviour for HbA1c; for example, a 30 minute displacement of MVPA, standing, sleep or SB into LIPA was associated with 2.33 (1.89, 2.77), 0.70 (0.31, 1.11), 0.63 (0.29, 1.00) and 0.42 (0.11, 0.78) mmol/mol higher HbA1c, respectively (Figure 6d). Note these displacement changes were observed in the age-sex-cohort models, but associations were attenuated after adjustment for covariates, most notably with the addition of physical limitations (Supplementary Tables 3-4, Models 2-3). While more time in SB was associated with higher HbA1c levels, with no impact of displacement between standing and sleeping (Figure 6a-c). The minimal daily behavioural change needed to observe a significant change in HbA1c was 3.8 minutes of MVPA displacing LIPA. A summary of all behavioural displacements across each outcome is provided in Supplementary Table 5.

### Sensitivity analyses

When regression models and reallocation plots were replicated using a four-part composition that combined standing and LIPA into one behaviour, results remained largely unchanged. However, there was a clearer and more consistent hierarchy of behaviours for all outcomes (Supplementary Figures 2-7); MVPA followed by LIPA then sleep were the most beneficial behaviours for positive cardiometabolic outcomes, with SB the most deleterious (Supplementary Table 6,7).

Compared to the complete cases sample (up to n=12,193), those missing one or more covariate (n=3,047) had lower HDL cholesterol (1.48±0.42 vs 1.57 ±0.47 mmol/L), lower HDL:total cholesterol ratio (3.61±1.22 vs 3.79±1.28), higher triglycerides (1.53±1.12 vs 1.47±1.02 mmol/L), and higher HbA1c (38.6±9.8 vs 37.9±8.6 mmol/mol) levels. However, adiposity measures were comparable and there was no different in movement behaviour compositions (Supplemental Table 8).

## DISCUSSION

In this large individual participant data analysis of over 15,000 participants, we examined cross-sectional associations between device-measured 24-hour movement behaviours and cardiometabolic health outcomes. Our findings revealed a clear hierarchy of favourable movement behaviours across the 24-hour day; MVPA was most strongly associated with healthier cardiometabolic outcomes. Using the mean 24-h behavioural composition as a starting point (7.7hrs sleeping, 10.4hrs SB, 3.1hrs standing, 1.5hrs LIPA, 1.3hrs MVPA), we observed theoretical benefits across all outcomes when as little as 4-12 minutes per day were reallocated into MVPA. Conversely, a greater proportion of time spent sedentary was detrimentally associated with all outcomes. More time spent standing was favourably associated with outcomes, although there were inconsistent – and often null – associations for LIPA. Associations between sleep and biomarkers were complex, with an unfavourable association when sleep replaced any time spent active (MVPA, LIPA, standing) and modest theoretical benefits when it replaced SB.

### Hypothesised mechanisms

The inflammatory, metabolic or vascular mechanisms through which MVPA contributes to improved cardiovascular health are well established^7, 8^. Our findings further suggest that even small changes in MVPA are associated with statistically significant and clinically meaningful cardiometabolic benefits. This builds on recent evidence reporting that small amounts of daily vigorous PA (accumulated in <2 minutes bouts) are associated with lower mortality, cancer and CVD risk^38, 39^. The acute benefits of standing on postprandial glucose response may partially explain the small but significant associations observed above^40, 41^. High muscle contractions involved in extended standing periods may also influence lipoprotein lipase activity, a key enzyme in glucose and lipid metabolism, and contribute to decreased inflammatory pathways^40, 41^. There were some positive associations of displacing SB or sleep into LIPA for BMI, however it was surprising to observe null or negative associations between LIPA and other cardiometabolic biomarkers. Given the inclusion of fast walking in MVPA, it is plausible that higher levels of slow walking (classified as LIPA) may be indicative of underlying health problems. Physical health limitations can contribute to greater levels of sedentary behaviour and less PA, and thus reverse causation may play a role^42^.

Mechanisms underlying association between insufficient sleep or too much sedentary behaviour and poor cardiometabolic health often focus on indirect factors that lead to weight gain or decreased energy expenditure^43^. However, chronic sleep deprivation has also been linked to the modification of gene expression and lipoproteins involved in inflammatory and cholesterol pathways^44, 45^. Our findings suggest that any theoretical cardiometabolic benefits from increased sleep – beyond the reference composition of 7.7.hours – are secondary to the direct physiological benefits of PA. However, it is unclear how the effects of displacing sleep and PA would differ in individuals with high levels of sleep deprivation. We hypothesise that individuals with insufficient sleep (i.e. <6 hours) may benefit from prioritising sleep over PA; the need for a more personalised approach to 24-hour behaviour is further discussed below.

### Comparison to existing evidence

Our study provides novel insights by distinguishing standing from ambulatory LIPA, and identifying the minimal theoretical displacements between behaviours required to observe statistical associations with cardiometabolic health outcomes. To our knowledge, this is the first study to suggest that more time spent standing may be more beneficial than LIPA for cardiometabolic outcomes. This must be interpreted with caution, given the likely inclusion of moderate-fast paced walking in MVPA rather than LIPA and the lack of context on the active or passive nature of the standing behaviour (e.g. resistance training, standing desk, waiting for a bus). Further research must investigate how context and the cognitive and musculoskeletal demands of standing and LIPA activities impact cardiometabolic health. Sensitivity analyses of the four-part composition is comparable to previous compositional studies, which have identified the benefits of MVPA and the detrimental consequences of SB for various health outcomes^17–19^. However, the studies reported inconsistent evidence regarding the role of sleep or LIPA activity on cardiometabolic outcomes, which may have been due to inadequate ascertainment of sleep using self-reported data^17^.

### Implications

Our findings have substantial implications from both research and clinical perspectives. First, they underscore the importance of MVPA across different adiposity and cardiometabolic biomarker outcomes. Our modelled reallocation suggest that population-level benefits can theoretically be observed after relatively short displacements of time (e.g. replacing other behaviour with 4 to 12 minutes of MVPA). However, it is crucial to examine if these effect sizes can be replicated in longitudinal observational or interventional studies using posture-based accelerometer data. Recently, there have been increased public recommendations on the ‘sit less, move more’ approach that highlights benefits of light-intensity activities for cardiovascular health^46^. The benefits of such activities may be more meaningful for mental health or musculoskeletal outcomes^47, 48^, rather than cardiometabolic outcome. Conversely, the findings here reaffirm the importance of the intensity of the activity that is replacing SB; our models suggest that replacing 30 min of SB with MVPA rather than LIPA result in substantially better cardiometabolic outcomes. It is crucial finding a balance between increasing time spent in higher intensity activities and decreasing time spent sedentary. For example, compositional analysis of hip-worn accelerometer data from the National Health and Nutrition Examinations Survey suggests comparable mortality risk between meeting US PA guidelines or by an additional 2.5 minutes of MVPA to ‘offset’ every 1hour of SB^18^. Therefore, optimal cardiometabolic outcomes can be achieved most efficiently if MVPA is specifically targeted.

Findings must be interpreted at the population level as the starting point for all reallocation plots is the mean sample composition, which has relatively high levels of sleep (7.7hrs/day), standing (3.1hrs/day) and MVPA (1.3hrs/day). Displacement into and away from MVPA did not demonstrate symmetrical associations with outcomes (Figures 1-6), and as introduced above, outcomes resulting from behavioural changes are likely to diverge depending on the initial starting profile. For example, previous investigation of dose-response associations between MVPA and cardiovascular outcomes have demonstrated steep risk reductions at low levels of MVPA, with benefits plateauing at higher MVPA volumes. Therefore, the theoretical benefits of displacing time in other behaviours into MVPA may be even larger in those who engage in little to no MVPA at baseline. This highlights an increasing need to identify personalised recommendations – or the “sweet spot”^49^ – based on an individual’s current 24-hour movement behaviours.

Modelling displacement of time between five key daily behaviours can inform design of more realistic lifestyle-based interventions and enable personalised behavioural changes. For example, interventions focusing on displacement between sleep, SB, standing and LIPA would likely require >1 hour of daily behavioural change to impact desired outcomes. This may have limited real-world plausibility compared to the potential impacts of displacing an additional ∼5 minutes in any other behaviour into MVPA.

### Strengths and limitations

Strengths of this study include the inclusion of 15,000+ participants from six cohorts and five countries to increase generalisability of our findings; the use of a thigh-mounted accelerometer wear position to sensitively capture postural changes; uniform ActiPASS processing of raw accelerometer data files; separation of standing from ambulatory LIPA; ascertainment of blood-based cardiometabolic biomarkers; and the complex compositional data analysis approach that simultaneously considered how time spent in different movement behaviours influences cardiometabolic outcomes.

There are some limitations that must be acknowledged. First, the data are cross-sectional, and therefore causality between movement behaviours and outcomes cannot be inferred. Recent mendelian randomisation of device-measured activity in UK Biobank suggest causal associations between MVPA and adiposity, with bidirectional associations between SB and adiposity^50^. Despite clear advances in the ActiPASS-based detection of activity intensity and sedentary behaviour, sleep time may have been overestimated as time spent in bed rather than biological sleep; nevertheless, previous work has suggested strong agreement between our sleep algorithm and polysomnography^33^. MVPA levels were very high in this cohort. This may be due to both specific cohort characteristics (e.g. high exercise sample in NES, manual occupation in DPhacto, etc.) or high levels of moderate activity classified as MVPA. Finally, there may be some residual confounding. Due to differences in measurement protocols between studies, some harmonised covariates had lower granularity than the original data collected (e.g. smoking, alcohol, medication use). We did not adjust the blood biomarker models for adiposity measures, to avoid overadjustment given that adiposity is likely to be on the causal pathway.

## CONCLUSION

This study provides novel evidence of the hierarchy of movement behaviours and their impact on cardiometabolic health markers. Findings emphasise a key public health message that positive cardiometabolic health outcomes can be most efficiently and feasibly achieved with small increases in MVPA. Standing – and for some outcomes LIPA – had positive associations with outcomes, although this was only observed after displacement of substantial amounts of time. SB was the sole behaviour with clear adverse associations with outcomes, regardless of duration. Compositional data analysis sheds novel insights on the complex interplay of 24-hour behaviours for cardiometabolic health outcomes. Taken together, our results suggest that prioritising a balance of more time in MVPA and less time in SB is the most efficient and effective way to improve and/or maintain good cardiometabolic health.

## Data Availability

All data produced in the present study are available upon reasonable request to the authors

## ACKNOWLEDGMENTS

The data on which this research is based were drawn from six observational studies in the Netherlands, United Kingdom, Australia, Denmark and Finland. We are grateful to all participants who provided the survey data.

## DATA AVAILABILITY STATEMENT

Access to data is not available directly from the authors of this manuscript. Access to cohort data may be available by contacting individual cohort and following their specific governance and access requirements.

## AUTHOR INFORMATION

ProPASS collaboration: Nidhi Gupta, Coen Stehouwer, Hans Savelberg, Bastiaan de Galan, Carla van de Kallen, Dick H.J. Thijssen

